# The lived experiences of people dying with frailty: a systematic review and thematic synthesis of qualitative studies exploring patients, relatives and professionals views

**DOI:** 10.1101/2024.10.28.24316304

**Authors:** Elizabeth Westhead, Daniel Stow, Bethany Kate Bareham, Felicity Dewhurst, Gemma Spiers, Lucy Robinson, Hannah O’Keefe, Fiona E Matthews, Barbara Hanratty

**Affiliations:** Population and Health Sciences Institute, Newcastle University, Newcastle upon Tyne, UK, NE2 4AX; Wolfson Institute of Population Health Queen Mary University of London, London E1 2AB

**Keywords:** Frailty, palliative care, end-of-life care, lived experience, care needs, qualitative

## Abstract

**Background:** Understanding the lived experiences of people dying with frailty is essential to develop models of care that are appropriate to meet the needs of this growing population.

**Aim:** Synthesise qualitative evidence on the experiences of people dying with frailty.

**Design:** Systematic review of qualitative literature and thematic synthesis. PROSPERO registration CRD42019141907.

**Data sources:** Fourteen electronic databases (CINAHL, Cochrane, Embase, EThOS, Google, Medline, NDLTD, NHS Evidence, NICE, Open grey, Psychinfo, SCIE, SCOPUS and Web of Science) searched from inception to May 2024. Studies were included if they reported on the lived experiences of people dying with frailty, and used an explicit measure of frailty for their sample. Quality was appraised using the Saini and Schlonsky checklist.

**Results:** Of 6,340 unique articles, 19 met inclusion criteria, describing the views of 138 people dying with frailty, 186 relatives /informal carers, and 240 professional caregivers. We identified three themes: ‘Identifying with frailty and dying’, ‘Emotional needs’, and ‘Support with daily living’. There was wide variation in people’s understanding of frailty, and of their proximity to death. Emotional responses to physical symptoms often had a greater impact on lived experiences than the symptoms themselves. People frequently reported a desire to live in the present, with priorities focused more on living than dying.

**Conclusion:** Approaches to palliative care for people dying with frailty should address emotional, as well as physical needs, and facilitate maintenance of existing daily routines. Ensuring that care planning accounts for individual understandings of frailty, and prognostic uncertainty may be particularly helpful.

**What is already known about the topic?:**

- Health and social care professionals and policy makers recognise the need to improve equity in palliative care provision for people dying with frailty
- A previous systematic review of quantitative literature highlighted the diverse range of physical, psychological and social needs of people dying with frailty

**What this paper adds?:**

- The narratives of people dying with frailty often focus on the emotional impact of physical symptoms, and fears around uncertainty
- People do not always recognise or identify with the concept of ‘frailty’, and unless actively dying, often express the desire to be supported to live independently for as long as possible
- Care providers express the need for holistic care, and voice frustration at service failures, including lack of time and personnel

**Implications for practice, theory or policy:**

- Services for people dying with frailty should be configured to be collaborative, flexible, holistic and responsive to changing needs
- Clinical training should emphasise the importance of monitoring patient needs, and both understanding and communicating uncertainty and unclear prognoses
- Future work and funding must now concentrate on developing and evaluating appropriate models of care

## INTRODUCTION

Improving provision of palliative care to ageing populations is an international priority. ^1^ The benefits of palliative approaches to care are clear: improved quality of life for people with malignant ^2^ and non-malignant diseases ^3^, greater likelihood of dying at home where preferred^4^, and fewer emergency department admissions.^5, 6^

The demographics of mortality are changing as global populations continue to age.^7^ People are increasingly dying later in life, often with greater medical complexity and multiple long-term conditions.^8^ The absolute number of people who die each year is also growing and these deaths are likely to be more protracted.^9^ Yet despite this growth in mortality, significant inequities in access to palliative services remain. People who experience greater socioeconomic disadvantage, ^10^ and have non-cancer diagnoses,^11^ are less likely to receive palliative care.^12, 13^

Another key population who face significant barriers to end-of-life care are older people; many of whom are living and dying with frailty. Frailty is a syndrome caused by age-related decline across multiple body systems, and which leads to a reduced capacity to respond to health stressors.^14^ In the UK, approximately 11% of people aged >65 years and up to 50% of those aged >85 years are likely to be frail.^15^ Older people with frailty may not access palliative care because of the challenges of prognosis in this population. However many people with frailty are dying with needs comparable to those of people dying with malignant diagnoses and organ failure.^16^ Policy makers and practitioners across healthcare recognise this challenge and are now giving greater attention to the palliative needs of older people with frailty.^17^

Efforts to identify optimal models of palliative care provision for ageing populations, including those with frailty, are ongoing.^18^ Understanding the lived experience of this population is fundamental to the design and implementation of appropriate models of care.^19^ To support user-informed service design, there is now a need to synthesise existing qualitative evidence about the lived experiences of people dying with frailty, their relatives, and carers.

### Aim

To systematically review and synthesise evidence regarding the lived experiences of people dying with frailty, from the perspective of patients, their relatives and carers.

## METHODS

A protocol was prospectively registered with PROSPERO (CRD42019141907), reporting follows updated PRISMA guidelines.^20^

### Search strategy

We searched CINAHL, Cochrane, Embase, EThOS, Google, Medline, NDLTD, NHS Evidence, NICE, Open grey, Psychinfo, SCIE, SCOPUS and Web of Science from inception until October 2017 using a search strategy developed in collaboration with an information scientist. We refreshed the searches in May 2024 using an additional qualitative research filter. Searches were supplemented with bibliographic screening and reference chaining. Title and abstract screening was undertaken by DS, with second screening by EW, GS, BB and FD. Final text inclusion was decided between DS, GS, BB, FD, and EW, with disagreements resolved in discussion with BH.

### Inclusion criteria

Design: qualitative studies reporting the views, perceptions, and experiences of being end of life with frailty from the perspectives of this population; family, friends and/or unpaid caregivers; or health and social care professionals.

Participants: people with frailty and nearing the end of life (with a definition provided within the article); receiving care in a palliative setting; or with stated terminal decline. We included studies that used a clear definition of frailty to define their end-of-life sample. Phenotypic, cumulative deficit or other operational definitions of frailty (integrating demographic, clinical, psychological and functional information) were all eligible.

Setting: studies reporting data from high income countries, as defined by the World Bank.

### Exclusion Criteria

Language: studies published in languages other than English were excluded unless translation was available.

Frailty definition: studies with no explicit definition or rationale for using frail to describe the sample (e.g. where ‘frail’ was used as a synonym for chronological age).

### Data extraction and quality appraisal

Data extraction was undertaken by EW and checked by BB. EW, LR and BB carried out quality appraisal using Saini and Schlonsky’s Quality Appraisal Checklist. Studies were not excluded on the basis of quality appraisal, as poor reporting is not necessarily indicative of badly conducted research.^21^ However, assessing quality prevents unreliable results from influencing review findings.^22^ Key limitations and comments on richness of presented findings are detailed for each study in Table 1 and summarised within our study descriptions..

**Table 1.** Detail of included studies.

| Citation | Country | Setting | Population | Design | Analytic method | Frailty definition | End of life definition | Total sample | N frail | Age (frail) | Gender (frail) | Key strengths and limitations including comment on richness from quality appraisal |
| --- | --- | --- | --- | --- | --- | --- | --- | --- | --- | --- | --- | --- |
| Bramley, L. 2016* | England | Participants recruited from wards at one hospital - most patients admitted following emergency admission from home | People moderately or severely frail, surprise question positive, initial reason for admission resolving (without severe cognitive impairment and/or on an active end of life care pathway) | Serial interview study | Bespoke analytic approach developed by lead author | Clinical Frailty scale (moderate and severely frail) | Surprise question positive | 16 frail older people<br>8 Relatives/<br>Carers (7 relatives, 1 close friend) | 16 | 85 (mean) | 10 (62.5%) female<br>6 (37.5%) male | Strengths: Transparent account of methods; longitudinal interviews across frailty trajectory; triangulation of insights from people with frailty and carers. Rich account of findings<br>Weaknesses: qualitative narrative unclear in places, with quotes not always obviously supportive |
| Bramley, L. et al. 2020* | England | Participants recruited from hospital wards for healthcare of older people. | Older people living with frailty who had recently been discharged from hospital | Serial interview study | Re-storied sequenced biographical narrative accounts for individual case studies, combined by cross case analysis | Clinical Frailty scale (moderate and severely frail) | Participants at very high risk of dying within the next few months | 16 frail older people<br>8 Relatives/<br>Carers (7 relatives, 1 close friend) | 16 | 85 (mean) | 10 (62.5%) female<br>6 (37.5%) male | Strengths: Transparent account of methods; longitudinal approach to data collection; triangulation of insights from people with frailty and carers are strengths.<br>Weaknesses: Thin account of findings, with intricacies left unexplored. |
| Carter, C 2022 | Canada | An urban academic Family Health Team in Ontario, Canada with 24 staff | Clinicians, caring for frail patients and having a scope of practice that includes EOL conversations. Patients over 65 with frailty who were medically stable. Care givers whom the patient's regular care depended on | Ethnographic study | Method of analysis not reported | Clinical Frailty Scale (mild to severely frail) | Patients considered to be approaching the end of life when a clinician anticipates that they may die in the next year | 20 clinicians<br>11 frail older adults<br>4 care partners | 11 |  |  | Strengths: Rich descriptions of setting, participants and analytical methods, triangulation of insights from healthcare professionals, people with frailty and observations within the setting. Weaknesses: demographic intricacies not explored in findings |
| Combes, S 2021 | England | A community based older persons' service run by a large urban hospice | Patients over 65 who were moderately to severely frail from a community based older person's service run by a large urban hospice | Individual or joint interviews in the participants home | Thematic analysis | Clinical Frailty Scale (moderately or severely frail) | Clinically assessed as likely to be in their last years of life | 10 frail elders<br>8 nominated family members | 10 | 85 (median) 71-95 range | 70% female | Strengths: rich description of analytic process. Weaknesses: Thin description of setting and participants. |
| Dodson, S 2024 | England | A district general hospital in the South West | Clinicians working in the District General Hospital | Individual interviews with clinicians | Thematic Analysis | Clinical Frailty Scale (severely frail) | Clinicians opinion that advance care planning for end of life would be appropriate | 7 clinicians | NA | NA | NA | Strengths: Thick description of the setting Weaknesses: Limited detail regarding of sample, no methods for triangulation have been described. |
| Emilsdottir, A.L. 2011 | Iceland | A long-term care nursing home located in the southwest part of Iceland - approximately 100 residents in total | RNs working in the nursing home | Ethnographic study | Interpretive phenomenology | Dependency based definition - entry to care homes in Iceland requires decision of pre admission committee - majority of residents in Icelandic nursing homes are very frail | Palliative care considered as commencing when residents move into the care home | 11 RNs<br>Unclear how many observations of other staff/residents | NA | NA | NA | Strengths: Mostly transparent account of methods. Triangulation of insights from RNs, older people with frailty and carers; ethnographic, interview and patient record data. Weaknesses: Account of findings offer some contextual detail, but exemplary quotes do not support this account. |
| Findlay, C. et al. 2017** | Scotland | Secondary analysis of a subset of interview data from Lloyd A, 2015 | People age 75 and over with frailty, MMSE of 24+ and without active/progressing cancer | Secondary data analysis | Thematic analysis | Clinical Frailty Scale (moderate or severely frail) | CFS moderate / severely frail mortality estimated 30% after 20 months | 11 frail older people<br>6 informal carers | 11 | 85 (mean) | 7 (64%) female<br>4 (36%) male | Strengths: Longitudinal interview data. Weaknesses: Thin account of findings, ideas unexplained and unevidenced; theme headings do not explain presented findings; quotes do not support findings. |
| Geiger, K et al. 2016 | Germany | GP practices in Germany | General practitioners recruited during a local training day and via Hannover Medical School's teaching practices network | Baseline and three follow up interviews per GP | Grounded theory | GPs are discussing their frail patients, and provide an estimate of their caseload who are frail | Questions to GPs focus on end-of-life care for frail patients | 14 GPs | NA | NA | NA | Strengths: Methods mostly transparent; longitudinal interview data; multiple researchers involved in analysis. Weaknesses: Thin account of findings relevant to this review; although unrelated intricacies were explored. |
| Kendall, M. et al. 2015** | Scotland | Existing literature (including a subset of interview data from Lloyd A, 2015) | Eight methodologically comparable, longitudinal, and multiperspective interview studies | Secondary data analysis | Thematic narrative analysis | Clinical Frailty Scale (moderate or severely frail) | Studies were all with patients nearing the (study described) end of life | 156 patients<br>114 caregivers<br>170 health professionals across multiple patient groups | 13 | 85 (mean) | 9 (69%) female<br>4 (31%) male | Strengths: Longitudinal interviews; multiple researchers involved in analysis; triangulation of perspectives of patients, family and practitioners are strengths. Weaknesses: Quantity of data seemed high for meaningful qualitative analysis; thin account of experiences of people with frailty, although comparison with experiences of different disease conditions provides additional insights. |
| Klindtworth, K. 2017 | Germany | Primary care practices in urban and rural areas within the region of Lower Saxony | People age ≥ 70 years with moderate and severe frailty, without dementia or progressing cancer | Longitudinal case study (up to 3 interviews - 6 month spacing) | Grounded theory | Clinical Frailty Scale (CFS), stage 6/7 | Study definition of people with advanced frailty towards the end of life | 31 people with frailty | 31 | 83 years (mean) | 19 female (61%)<br>12 male (39%) | Strengths: Transparent account of methods; longitudinal data collection. Thick description of findings. |
| Lloyd, A et al, 2020** | England | Community dwelling participants recruited from a medical day hospital | People age 75 and over with frailty, MMSE of 24+ and without active/progressing cancer | Individual or joint interviews (with the informal carer), of between 35 and 120 minutes were carried out up to three times over a mean period of 18 months | Voice-centred relational method (VCRM) | Clinical Frailty Scale (moderate or severely frail) | Study defined - discussions and narratives around end of life | 13 frail older people<br>13 linked carers<br>8 professional carers | 13 | age range 76-92 | Unclear | Strengths: Transparent account of methods; longitudinal data collection; triangulating experiences of people with frailty and carers; rich contextual detail of individual cases. Weaknesses: Thin description of overall findings beyond individual experiences |
| Lloyd, A Et al. 2016** | Scotland | Existing literature | Four theses on end of life experiences of people dying with bowel cancer, glioma, liver failure, and frailty | Secondary data analysis | Three stage analytic process involving 1) thematic analysis of interviews in each thesis focussing on palliative issues, 2) Exploring differences between older and younger age groups in each thesis, 3) Synthesis of findings from all four theses | Clinical Frailty Scale (moderate or severely frail) | The four primary sources explored "end of life experiences" - each had their own definitions not further explained in the secondary data analysis | 65 people at end of life across multiple diagnostic groups | 13 | 85 (mean) | 9 (69%) female<br>4 (31%) male | Strengths: Different insights triangulated. Weaknesses: Reported findings not supported with quotes; thin account of study findings. |
| Lloyd, A. 2015** | Scotland | Participants recruited from an outpatient/day hospital | People age 75 and over with frailty, MMSE of 24+ and without active/progressing cancer | Serial interview study | Voice Centred Relational Method (VCRM) | Recruitment definition "CFS severely or moderately frail and receiving 7 or more hours per week of formal or informal care" | CFS moderate / severely frail mortality estimated 30% after 20 months | 13 people with frailty | 13 | 85 (mean) | 9 (69%) female<br>4 (31%) male | Strengths: Transparent account of methods; longitudinal interviews; triangulation of insights from people with frailty and carers; rich narrative of findings. |
| Nicholson, C et al, 2012 | UK | Community dwelling older adults recruited via intermediate care team | Community dwelling older adults with frailty | Serial interview study | Biographic Narrative Interpretative Method | People of advancing age, unable to carry out independent activities of daily living (Lawton & Brody, 1969) and considered to be vulnerable to physical decline | participants had multiple and accumulating health needs, and became increasingly dependent and frail as the study progressed (although no participants died during the 17 month duration of the study, one participant died 8 months after study end) | 17 people with frailty | 17 | range 86-102 | 12 (71%) female<br>5 (29%) male | Strengths: Mostly transparent account of methods; longitudinal data collection; triangulating data from multiple sources; researchers from different backgrounds contributing to analysis.<br>Weaknesses: approach to analysis unclear; contextual detail is provided in account of findings; although intricacies are not considered and it is not always clear whether ideas are grounded in participant experiences. |
| Overbeek, A et al, 2019 | Netherlands | 16 residential care homes participating in an RCT of ACP | Relatives of people with frailty age ≥75 | Interviews with bereaved relatives: Part of an RCT - residents and relatives offered ACP (intervention) or care as usual (control) | Thematic framework analysis | Tilburg Frailty Index Score ≥5 | All interviews with bereaved relatives | 201 people with frailty in RCT<br>39 relatives participated in qualitative study | NA | 89 (mean) | 30 (75%) female<br>9 (25%) male | Weaknesses: Large delay between event of interest and data collection; thin description of findings provided; few negative findings presented. |
| Piers, R D. et al, 2013 | Belgium | Three different geriatric care settings: (1) nursing homes, (2) an acute geriatric ward, medical oncology ward and palliative care unit of the Ghent University Hospital and (3) home services for elderly staying at home but requiring help for activities of daily living | Patients aged 70 years or older who healthcare providers believed to have a limited life expectancy | Single interviews | Thematic analysis | Lunney end of life trajectories | Surprise question positive | 21 people with terminal cancer<br>17 people with frailty or end stage organ failure | ~17 | (total sample) median 81 years, range 71-104 | 26 (68%) female<br>12 (32%) male | Strengths: Mostly transparent account of methods; triangulation of experiences of older people with different end of life circumstances.<br>Weaknesses: Thin description of findings in relation to experiences of frailty. |
| Skillbeck, J.K 2014 | England | Community dwelling older adults recruited via community matron services in a post-industrial city in the North of England | older people with complex health problems living at home receiving care from health and social care workers | Ethnography/<br>Case study design | Thematic narrative analysis within and across cases | Community matron opinion based on researcher provided description as vulnerable; fragile; dependent; at risk (taken from Browne and Markle-Reid 2003). | 6 of 10 participants died during study (and within 1 year of first observation) | 10 cases: a case was defined as an older person, a community matron, and sometimes a significant other, such as, daughter, neighbour | 10 | median 84 years, range 77-91 | 7 (70%) female<br>3 (30%) male | Strengths: Transparent account; ethnographic and interview data; triangulates insights from OPwF, SO and practitioner; contextually grounded findings.<br>Weaknesses: demographic intricacies unexplored in findings. |
| Stiel, S, 2019*** | Germany | GP practices in Germany | General practitioners recruited during a local training day and via Hannover Medical School's teaching practices network | Baseline and three follow up interviews per GP | Grounded Theory | GPs are discussing their frail patients, and provide an estimate of their caseload who are frail | Questions to GPs focus on end-of-life care for frail patients | 14 GPs | 0 | NA | NA | Strengths: Longitudinal interview data, thick description of setting and context.<br>Weaknesses: Approach to data extraction from original study unclear. |
| Stuart, K, 2019 | England | Case reviews identified from mortality register of one NHS trust | People age 75 and over with frailty, without progressing cancer, living in the community at time of death | Case reviews of deceased frail older patients | Connelly and Clandinin (2000) commonplaces framework | 7-8 on clinical frailty scale at the time of care | All people with frailty deceased | 3 nurses<br>4 occupational therapists<br>3 physiotherapists | NA | 89.6 (mean) | 2 male<br>1 female | Strengths: Transparent account of methods, mostly appropriate to study aims;<br>Weaknesses: practitioners interviewed three months post-care of one patient, raising concerns of recall issues; medics' voice missing from care team of study; descriptive account of findings with little exploration of intricacies. |

\* Data from Bramley 2016, \*\*Data from Lloyd 2015, \*\*\*Data from Geiger 2016

### Analysis

A thematic synthesis of included studies was conducted.^23^ Methods for synthesis were based on Braun and Clarke’s principles of thematic analysis.^24^ The review team familiarised themselves with findings of each study during full text screening. The lead author listed ideas and potential codes from primary study findings during this phase. Compiled codes were comparable to second- and third-order constructs described in meta-ethnography. Second-order constructs are interpretations and themes derived from primary data, specified by authors of included studies. Third-order constructs are interpretations identified by the review team that further explain findings within and across primary studies^25^. To examine experiences across the disease process, papers were broadly coded as to their focus within the disease trajectory; for example, frailty severity and symptom management was coded separately from papers exploring issues of deterioration, end-of-life care and bereavement. Recurring codes, explaining findings across studies, were developed into a candidate framework of themes that explained the lived experiences of older people dying with frailty.

NVivo (version 11) was used for data management. The lead reviewer recorded analytical notes during this process, detailing explanations and patterns within each theme. The thematic framework was further refined to ensure that it reflected views and experiences conveyed across included studies, and defined to form the theme descriptions presented as our findings. Excerpts from included studies (supporting quotes from study participants and extracts from study authors’ narratives) were identified to present as examples. Developing themes were discussed amongst the multidisciplinary research team to inform data interpretations, which included providers of both generalist (BH) and specialist palliative care (FD, LR), and academics with expertise in health psychology (BB), methodological (BB) and topic (DS, FD, LR) experts. The derivation of themes was predominantly inductive, although there was a deductive component as the process was framed by consideration of disease trajectory. Multiple studies reported on information drawn from the same patient sample, and one study reported secondary analyses of existing data. In the summary of studies, demographic information from these was only recorded for the original source.

### Theoretical position

This qualitative review and synthesis was conducted from a critical realist perspective; assuming a real world exists independently of perceptions, but understanding of this world is constructed from individual perspectives. By examining the experiences of older people with frailty, their family and carers, and practitioners, we can gain a more complete understanding of their support needs by triangulating these different perspectives. We drew on Bradshaw’s Taxonomy of Social Needs in our analysis to consider different types of support needs evidenced in included studies.

## RESULTS

### Eligible studies

Searches identified 6,340 unique results, of which 6,169 were excluded following title and abstract review, leaving 171 studies to be assessed for full eligibility (Figure 1).

**Figure 1.**
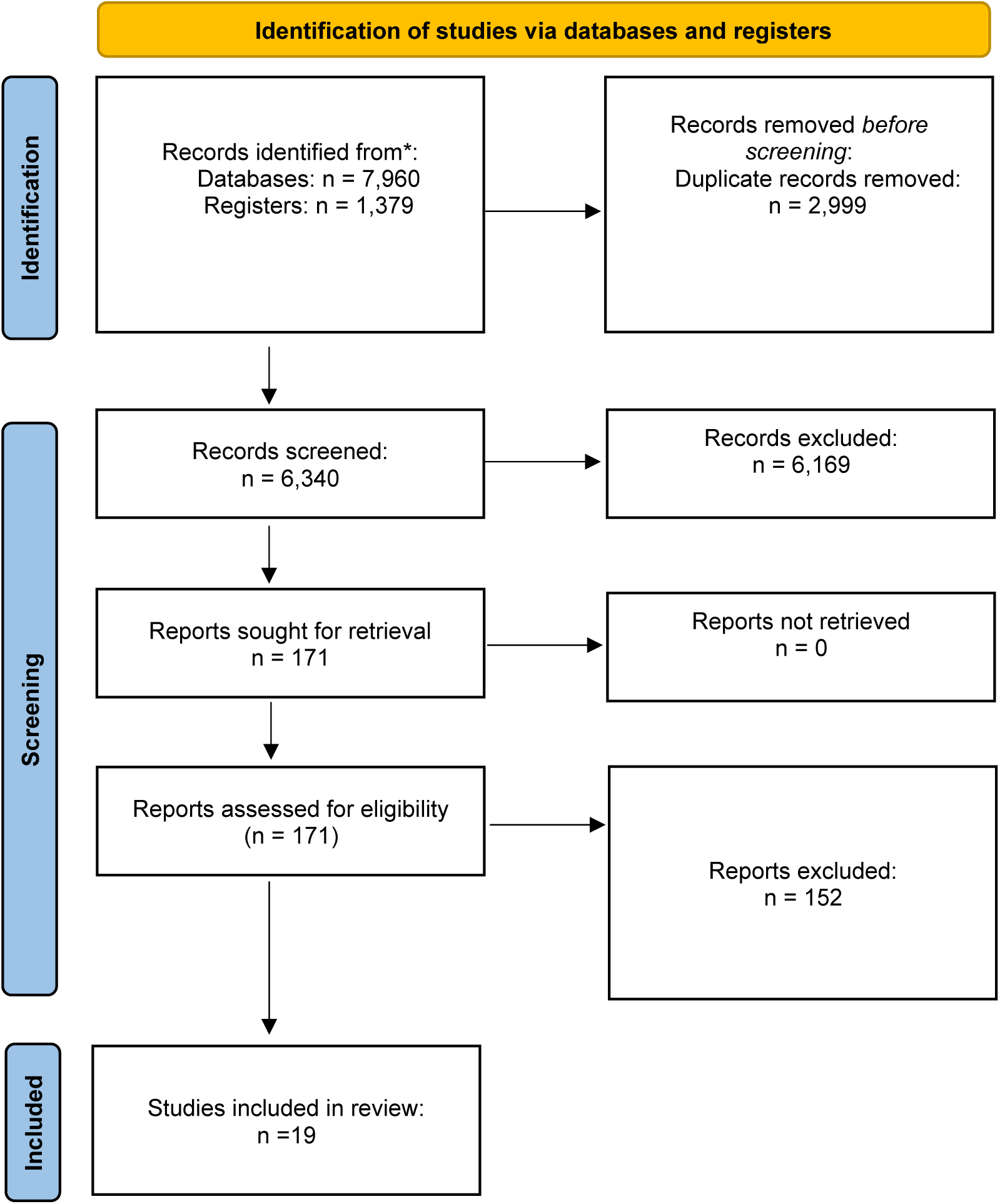
Prisma flowchart of included/excluded studies.

### Characteristics of the included studies

Nineteen papers met eligibility criteria, reporting thirteen unique studies published 2011-2024. Two studies presented analysis of data from Bramley, L, 2016 (Bramley, L. 2016* Bramley, L. et al. 2020*), two studies analysis of data from Geiger (Geiger, k et al. 2016, Stiel, S et al. 2019) and five studies analysed data from Lloyd, A. 2015 (Findlay, C. et al. 2017** 2016 Kendall, M. et al. 2015**, Lloyd, A et al, 2020**, Lloyd, A Et al. 2016**,Lloyd, A. 2015**). Table 1 contains details of all included studies and participant demographics. Unique demographic information was available for 13 patient samples. Included studies were from the United Kingdom ^13, 26–35^, Germany ^36–38^, Belgium ^39^, Iceland ^40^, the Netherlands ^41^ and Canada. ^42^ The studies’ foci were people with frailty (n=138), relatives/informal carers (n=186) and healthcare professionals (n=240). Fifteen papers were published peer-reviewed articles, and four were doctoral theses.

### Quality appraisal

The main quality limitations related to a lack of transparency in reporting, particularly limited contextual detail and depth^35^ ^13, 27, 29–31, 36, 39, 41, 43^; quotes unsupportive of reported findings ^28, 40, 43^; and a time delay between participants’ experience of the phenomenon of interest, and data collection ^34, 41^. Narrative appraisals for each included study are provided in Table 1.

### Main Findings

We identified three overarching analytical themes: “Managing the physical and emotional impact of frailty”, “Continuity and autonomy in day-to-day life”, and “Identifying with frailty and dying”. Within each theme we draw on the experiences of people with frailty, relatives and domiciliary caregivers, and clinicians.

### Managing the physical and emotional impact of frailty

Physical symptoms of people with frailty varied greatly on an individual basis, and from day-to-day; ^26, 28, 32, 33^ and included pain, nausea, breathlessness, weakness, fatigue, limited mobility and insomnia. ^26–31, 36, 40^ Symptoms affected their ability to engage in valued hobbies and interests, and social opportunities; ^26, 28, 43^ as exemplified by the experiences of an older man in Lloyd and colleagues’:

> ‘Mr Mackie struggled further with deteriorating eyesight which was a significant issue because of the impact on his routine. “But his loss of reading through his eyesight that’s…” “It’s a big thing, mm-hmm. Because he used to go through three books a week, you know, and his Observer, his this and that” Having faced the loss of other activities, “I had a fair mixture of hobbies and so on that it always kept me going. I feel terrible, stuck now” being unable to read rendered him unoccupied and bored and frustrated in the day. “You get bored day after day just sitting here you know” (Older man with frailty, caregivers, and analytical narrative) ^26^

Some adapted to their changing abilities by engaging in their hobbies and interests by different means: ^26, 28, 43^

> “’Despite barriers including diminishing eyesight and hearing, participants strived to pursue the things they enjoyed by adapting them to suit their changing health needs. Where [older man with frailty] had once enjoyed going to his allotment and digging his vegetable patch, he now liked to ‘play around with his pots’ outside his back door.’ (Analytical narrative)^28^

However, it was not always possible for people with frailty to find alternative ways to satisfy their interests that met their abilities.

Reduced mobility was the condition highlighted by people with frailty as most problematic, as it impacted their independence, and their ability to leave the house. ^27, 34^ They found themselves increasingly socially isolated, and could become lonely ^26, 28, 33, 37^ with visits from caregivers often their only source of social interaction: ^28, 32, 33^

> ‘For [people with frailty] that lived alone, loneliness was a real problem. Most participants had family or friends that visited them regularly, but this did nothing to compensate for the long hours they spent in their homes unable to leave.’ (analytical narrative)^28^

Cognitive decline was a particular worry for people with frailty, causing further struggles to engaging in enjoyable activities, and fears they would lose their sense of self:^26, 28, 31^

> ‘[older people with frailty] were worried about their ‘mind going’. Stanley accepted that old age was something that he was unable to control, but he was worried about ‘losing his mind’. “Yes it is one of them things; we all get older. As long as my mind doesn’t go, that’s the big worry…”’ (Older man with frailty and analytical narrative)^28^

Older people described living with their declining abilities as a ‘restricted existence’, and their lives ‘closing in’: ^28, 31, 33, 43^

> “I think the most worrying thing is visualising the way our lives are going to contract… and the loss of freedom to do things in our lives in our own way.” (Older man) ^28^

Poor emotional wellbeing was emphasised in accounts of people living with frailty ^26, 33^ and in the concerns of their family members and care providers regarding their relative/patient ^26, 32, 33, 40^. Many people with frailty experienced symptoms of depression and/or anxiety relating to living with their condition^26–28, 34^. Worries and low mood linked to their physical/cognitive decline, pain, social isolation, reduced abilities, concerns about financing future care, further health issues, hospitalisations, entering residential care, and the prospect of death and dying, ^26–28, 32–34, 37, 39, 40, 43^ as Skilbeck^33^ describes:

> ‘It was not always possible for the older participants to adopt a positive outlook to their current situation, especially where there was deterioration in their condition. [Older woman with frailty] started to become very tearful during the observations and interviews towards the end of the study as she was becoming increasingly overwhelmed and distressed by the changes to her health and the ensuing loneliness. She often mentioned that she did not know what was wrong with her, “I don’t know why I am like this.” At one point [older woman with frailty]’s conversation started to be dominated by her anger and frustration at her increasing breathlessness as it indicated to her that her condition was getting worse. She was beginning to feel fed up and she did not know how to improve her situation.’ (Older women with frailty and analytical narrative)^33^

Mental health symptoms associated with frailty could be debilitating, and an additional barrier to basic activities such as leaving the house; ^26, 28^ as exemplified in this case of an older woman in Bramley’s study^28^:

> ‘On numerous occasions [older woman with frailty] had experienced breathlessness which had led to panic. These episodes of panic had in turn left her feeling as though she could not attempt to do things that she had previously taken for granted. “I am still out of breath… very much so… I am too frightened to go outside. I mean, I stood outside the other day with two carers with me, and I just went rigid. I panicked and I just stood in the street and cried you know…” [Older woman with frailty]’s panic attacks had meant that she no longer wanted to leave her house. This meant that she had missed her desperately needed follow-up appointment at the hospital with a specialist, at the time she probably needed it the most.’ (70 year old women with moderate-severe frailty, and analytical narrative)^28^

Palliative care professionals prioritised the emotional needs of patients with frailty alongside other symptoms, such as pain.^40^ However, clinicians with a broader remit beyond palliation, such as general practitioners, struggled to address these wider needs for wellbeing within appointments: ^26, 36, 38^

> “GPs don’t have enough time, I mean you give [people] lots of time and in a home visit package of care assessment, different assessments, different needs. And then you step back because you’ve done your medical management bit as much as you know that there are social, spiritual, emotional needs ah, you haven’t the time to invest in that” (General practitioner)^13^

### Continuity and autonomy in day-to-day life

People with frailty were keen to maintain continuity in their day-to-day routines. ^26–28, 31–33, 39, 42^ Grounding themselves in the present through routine distracted from worries about the future; ensured they maximised their enjoyment of their remaining time; and helped them retain a sense of autonomy and independence.^26–28, 30, 33, 39^ As older people’s abilities declined, many had to adapt their day-to-day activities to manage their frailty; prioritising those they felt to be most important, and doing more on days when they had more energy: ^26, 28, 32, 33, 37^

> ‘In order to manage the effects of physical ill-health and impairment, maintaining a daily routine was important. Engaging with such routines appeared to be significant as it enabled [people with frailty] to sustain their sense of self as an independent person through the control of habitual routines. I observed how patterns of the day were structured and adhered to. For example, [older man with frailty] routinely had an early lunch and then retired to his bedroom so that he could lie down and watch television; this also conserved his energy for his bedroom routine. Similarly, [older woman with frailty] would lie down after breakfast to read and then after lunch would watch a series of television programmes up to bedtime. When routines were easily accommodated the impairment seemed to go out of focus. (analytical narrative).^33^

People with frailty valued their autonomy. ^26, 33, 37^ Almost all prioritised remaining in their own home: this finding cut across cultures. ^26–28, 31–33, 36, 37, 43^ They avoided hospital admissions until they felt their health state was intolerable, and moving into residential care was considered a last resort when they could no longer be supported to live at home.^26–28, 31, 33, 37, 43^

> “The warden was saying when we came back from the hospital you know “you wanting to go into a home” but we don’t want to do that. As long as we can stay in the flat, as long as we can manage” “It’s always going to be a last resort, I mean neither he nor I are stupid and I mean we know that place could come one day but we hope we don’t have to do that eh no?” “No it’s a last resort” “A last resort” (Discussion between older couple with frailty). ^13^

Family members recognised and shared this stance;^26^ and general practitioners strived to maintain people with frailty in their homes. ^36, 40^ People with frailty who had had to leave their homes for care described distress over their loss of autonomy.^26, 34^

People with frailty were often reluctant to accept support from family or formal caregivers to aid them in day-to-day living; or adopt adaptive equipment to help with mobility issues. ^26, 28, 32, 33, 37^ They held concerns about becoming a burden ^26, 28, 33, 37, 43^ and the concept of losing their independence. ^26–28^ Those who accepted support only did so after they were no longer able to carry out the activities they had been struggling with; ^26, 43^ but conveyed that they valued this support, as described by Lloyd regarding the experiences of an older couple with frailty^13^:

> ‘Although they seemed to have come to terms with their circumstances, [older woman with frailty] described the inherent struggle in the process of getting to this stage “We didn’t want it [support from formal caregiver] and [husband] particularly didn’t want it but it was a big step to do that and we accept now we can’t do without the help so don’t be proud, take the help and make your life easier and you’ve got time to sit and enjoy each other’s company. Accept all the help that you’re given”’ (Older woman with frailty). ^13^

However, people with frailty sometimes felt that necessary support with more intimate aspects of care, such as toileting, was infantilising. ^26, 43^

This ‘scaffolding’ of their day-to-day lives was important for people with frailty to maintain their standard of living. ^26, 28, 33, 36, 37, 40, 43^ Domiciliary carers could support the person with frailty to remain in their home, and continue their lifestyle, ^27, 28^ as exemplified by the experiences of an older woman described by Lloyd^13^:

> “Day-to-day things like cooking, you manage that?” “Ocht no bother” “and cleaning?” “I have two people come in on a Monday and two people come in on Thursday privately and they come in and they do all my hoovering and dusting for me and [the gardener] of course comes in on a Monday for four hours and does the garden so I’m alright” “You’re grand. And you manage to have a bath or shower” “Well, yes eh because I had a shower this morning because [carer] comes in and on a Tuesday I have a shower” (Interviewer and older woman with frailty)^13^

Inconsistent or inflexible timings for this care provision disrupted their routines and their day became consumed by awaiting care rather than engaging in routine activities and living well: ^26, 28, 32, 33, 43^

> ‘[Older woman with frailty] was frustrated with the regimented nature of the home care system. For her, this often meant that she felt rushed to get out of bed in the morning or was left in bed too long, sometimes until mid-morning. On a daily basis, she had no idea what time the carers would arrive. “I say to them in a morning when they are rushing me, “Don’t rush me, please”, cause I get an hour in the morning, and I get half an hour at tea time and I get a quarter of an hour to put me to bed. Well, that is the only thing I don’t like, and I will be honest with you sometimes they put me to bed at eight o’clock, but they are not supposed to put me to bed ‘til half past eight, twenty to nine”’ (Older woman with frailty and analytical narrative, Bramley)^28^

Not all accepting of care had a support network capable of meeting their needs, or access to professional care, and in such cases people with frailty could struggle to manage their self-care routines. Instances were described by Bramley where the person’s health deteriorated rapidly, and their care needs increased. ^28^ Care services could find it difficult to secure adequate care in a timely manner, leaving them without appropriate support with their day-to-day living^28^.

### Identifying with frailty and dying

For people with frailty, their level of identification with their condition and prognosis influenced their experiences of living and dying with frailty; and their needs regarding how discussions about future care were navigated by clinicians.

Whilst people living with frailty recognised their poor health state, most did not view themselves as ‘frail’: ^28, 32, 33, 37^

> ‘Although all the older people were able to describe the physical and cognitive characteristics that in their opinion indicated frailty the majority of them did not consider themselves to be frail. In fact, on the whole they vehemently resisted the label when asked if they considered themselves to be frail.’ (analytical narrative)^33^

Their lay definition of frailty reflected an inability to manage basic activities of daily living, and a total dependence on support from others; which often did not reflect their own health state.^26, 33^ Many perceived their declining health to be a natural part of getting older ^26, 28, 33, 37, 43^ or were still awaiting to be diagnosed with a specific condition that would qualify their need for help and care from others. ^26, 31, 33^ Others associated ‘being frail’ with being ‘very old’; and when they did not consider themselves to be ‘very old’, neither did they recognise themselves to be frail.^26^ Some perceived their declining health was temporary, and expected to recover. ^26, 33^ People with frailty and their practitioners often had contradicting understandings of the person’s health state and prognosis; as exemplified in a case described by Lloyd^26^:

> ‘That his (the patient’s) physical decline did not mean the same thing to him as it did to others was highlighted by the GP. “I mean he’s (the patient’s) gradually become more frail and less able. He doesn’t really see that. He considered undiagnosed viruses” (narrative regarding 91yo man and his GP)^26^

Clinicians cited physical, social or emotional vulnerability relating to patients’ physical decline as evidence for their frailty. Such discordant perspectives regarding the cause of physical decline could lead to frustration on the part of the person with frailty during consultations^35^ ^26, 33^.

People who had come to recognise and accept their frailty, and that a complete recovery was unlikely for them, did so following sustained and significant decline: ^26, 28^

> ‘Miss Pegg remained accepting of physical decline and no longer clearly hoped for improvement “It’s just a general decline, you know, … I just feel I’m not doing as well as I used to, you know.” Resigned that a full return to previous function wasn’t going to happen, she envisaged a gradual, inevitable decline “Well, I might get a bit better but, you know, every time I go in and something happens you never quite get back to where you were” ^26^

People with frailty recognised their trajectory towards death as part of their life stage, and made practical arrangements in preparation; including wills and funeral plans. However, when not yet severely limited by their health state, people with frailty usually did not recognise death or dying to be an imminent eventuality. ^26, 28, 30, 33, 39, 42^ This was exemplified in Lloyd’s findings regarding an older couple:

> ‘Looking to the future the couple explicitly talked of their lives nearing their end. Robert: “Well we’re both in our 80s, well I’m 85 and Kathleen is 84, we don’t really have statistically great expectations of life you see, sorry but you have to face it.” Yet death remained abstract and distant and they expected their lives to continue for the next few years.’ (Analytical narrative/quote from older man, aged 85 years)^26^

Many people with frailty were distressed by the idea of dying, and preferred not to dwell on this eventuality^27^. It was evident that few had received a direct indication from their clinicians that they were approaching the end of their life.^26, 31, 32^

> ‘No frail patient described having had any discussions about planning for future deterioration or the end of life. Instead, they took a more reactive ‘deal with it when it happens’ approach […] despite understanding their mortality, death remained abstract for them’ (Analytical narrative)^26^

The uncertain trajectory of frailty, and their patients’ lack of recognition that they were dying, meant that clinicians were reluctant to adopt palliation, or initiate advanced care planning with people with frailty^35^. Clinicians reported that it was difficult to engage such patients in meaningful discussion regarding wishes for future care when their understanding of their prognosis was so discordant with their own.^30–32^ End-of-life care planning was often only broached with people with frailty in emergency situations when receiving care for acute health events.^26, 28^ People with frailty, and their relatives, described the confusion and distress they felt during such incidents, as they learned of their prognosis for the first time in already chaotic and upsetting circumstances: ^26, 28^

> ‘Two participants described being involved in very forthright conversations with healthcare professionals regarding their end of life. In both of these cases, participants recognised that they had been identified as dying or could die imminently as a result of their current condition. As a result, they had been engaged in discussions relating to this while they were in hospital. Both had been part of crisis discussions with medical staff around ‘do not attempt cardio pulmonary resuscitation’ and withdrawal of treatment. For both women, this was the first indication that they may have reached the end of their lives and may not survive much longer. As a result, both women had been left with unanswered questions and feelings of helplessness about their future in or out of hospital’ (Analytic narrative)^28^

Where a palliative approach to care had been initiated with a person with frailty, clinicians described fewer challenges in discussing end of life care with these patients. ^36, 38, 40^

People with frailty who had experienced physical/cognitive decline to the point of losing hope of any potential recovery from their current state were often accepting, and even welcoming of death, as an end to their struggles and a life that they now felt lacked meaning ^26, 32, 33, 39, 43^ for example:

> ‘Mr Hughes was negative describing a futile existence: “You say to yourself, ‘Well is it worth it, all this nonsense?’” By now Mr Hughes felt his life was intolerable. “You feel you want to walk out there and walk under a bus. I get so frustrated now, you get angry…” Gillian [daughter] repeatedly told me how her father had had enough of existence. “I mean he will come out with things at times, you know ‘I’ve had enough’” (Analytical narrative/older man with frailty/daughter)^26^

Some of the people with frailty who accepted their current trajectory towards death described advanced care planning (ACP) conversations with their GP, facilitating them to consider a palliative, rather than curative, expectation for care. ^26^ For these individuals, ACP conversations brought comfort, resolving their sense of uncertainty about the future, and helping them feel that their autonomy would be respected. ^26, 41^

## DISCUSSION

### Main findings of the study

This systematic review reports the experiences of 123 people dying with frailty, 186 relatives or informal carers, and 240 professional caregivers: it explores the all-encompassing effects of frailty and how it in-discriminatorily impacts on all aspects of patients and their relatives’ lives. In turn patients, relatives and personal and professional carers make clear that any effective palliative intervention must be holistic to reflect their needs.

Our findings highlight the need for person-centred individualised care within this population. Patient autonomy is important, and key to ensuring a sense of control. Individuals and carers described their progressive frailty as a cycle of unrelenting loss and ambiguity. Individuals strove to maintain miautonomy in their decision-making even when increasing disability compromised autonomy in their daily activities. It was often individuals’ choice to stay in their own homes. However, care and support frequently were not responsive or appropriate enough to support these preferences. Relatives’ choices were often at odds with that of the patient, where worry for their loved one meant they encouraged choices such as moving in with relatives or institutionalised care. This may exacerbate patients’ loss of autonomy and introduce feelings of “being a burden”. Support for patients and relatives, both individually and as a family unit, is vital to help people adjust to such progressive loss and maintain a sense of control. Similar experiences have been reported by other patients with neuromuscular disease (NMD) where trajectories of illness and rates of deterioration may be difficult to predict.^44^

Knowledge of the trajectory of frailty was lacking amongst patients, relatives and healthcare professionals alike. Triggers of deterioration and key points of decline were often missed and as such realistic prognostication failed to occur. This is also recognised in populations with NMD^44^ where it has been noted that specific factors should trigger palliative care input. For people with frailty, the trajectory of decline may be gradual and such triggers may not be obvious. End of life for this population may not be linear or a smooth continuum. Consequently, advance care planning assumes a greater importance.

Effective communication and realistic prognostication are key to appropriate advance care planning, realisation of end-of-life care wishes and as a result respecting patient autonomy.^45^ Advance care planning and good palliative care are synergistic. Within our review, people who had been supported to plan ahead and were able to access palliative care services earlier described a calm end-of-life experience, whilst relatives experienced less prolonged grief in the bereavement phase. Encouraging healthcare professionals to initiate advance care planning early in the trajectory of frailty can normalise such conversations^46^. Early and effective communication may help individuals plan for deterioration and set individual ceilings of care accordingly. Such communication should ideally be facilitated by a single advocate who is integrated within a multi-disciplinary team and who has experience of caring for people with frailty. Anticipating and discussing intervention options to manage acute crises is critical. Effective communication between patients, family members and healthcare professionals from different care settings is fundamental to the success of advance care planning, without which decisions may not be adhered to.

In line with previous work ^16, 47^, this review emphasises the importance of individualised care. It is also imperative to explore the range of different preferences regarding information needs, interventions and services. Individuals will have varying attitudes to; decision-making, the future, dying and practices post-mortem and these should be respected. The way in which people living with frailty formulate their end of life care preferences, and how these may change over time is currently being explored.^48^ Any framework for improving palliative care for people with frailty and their carers should take account this into account.

This review suggests that relatives are affected by frailty in similar ways to those living with it. Carers experience frustrations, confinement and isolation, whilst the sense of burden grows as their relatives’ condition deteriorates. Carers also report carrying anticipatory grief alongside guilt at not being able to do more. These experiences are compounded by the fact that relatives may themselves be frail: a scenario that will become increasingly problematic as populations age. The importance of caring for the “family unit” is highlighted and ongoing input after the death of the primary patient is likely to be required. This may include both bereavement (exacerbated by societies ambivalence to a perceived “inevitable” death) support and holistic care when the carer becomes the now frail patient.

Healthcare professionals can struggle to manage frail patients, particularly those who realise that too many things are going wrong for the medical model of care to be sufficient. Any lack of palliative care knowledge amongst generalist and specific frailty teams may be met by a similar lack of frailty knowledge amongst palliative care teams. The answer is likely to lie in multidisciplinary teams and joint working between different specialist groups, both challenging to organise. Our findings support the holistic model of palliative care to accommodate the needs of frail individuals. We explored effective exposure to (or lack of) palliative care and considered when and how palliative care should be delivered. Effective strategies push aside the medical model of care and embrace holistic caring, highlighting the importance of social care specifically and the multi-professional team in general. In an ideal situation, exposure to, and experience of, palliative care could be normalised across the trajectory of frailty, alongside advance care planning. Prompt and recurrent access has been previously suggested as an effective strategy.^6^ If palliative care and hospice visits could become part of routine care, fear and stigma may be able to give way to services and places that offer proactive solutions and excellent care. Access to such support would enable patients and carers to have a better overall experience of living and dying with frailty, helping mitigate against the inevitable loss and uncertainty felt as a result of the disease.

### Strengths

This review includes extensive international qualitative evidence detailing individuals’ experiences. Findings give a voice to patients, relatives and healthcare professionals. To ensure that the results from this review remained relevant to all, our stakeholders who contributed to the study development included palliative care generalists and specialists. The depth of available data is also a strength: the complexity and completeness of accounts contained with the four themes in particular facilitated a rich synthesis.

### Limitations

A criterion of this review was the need for an explicit definition of frailty. This enabled our synthesis to describe the needs of a clear and precise population. However, this approach also excluded studies where frailty might have been described in different ways. For example, we did not include studies where participants were described as having a low performance status, a label used by oncologists and palliative care physicians. As such, our synthesis is limited to populations who are explicitly described as frail. We did not exclude studies by country. While this broadens the findings to a more international focus, the papers therefore reflect experiences across a disparate set of healthcare systems. Finally, most of our included studies drew data from people living in their own homes. Populations living in alternative settings, such as care homes, are therefore underrepresented in our synthesis. More research is required to understand their needs, and how such needs may differ from people living in individual households.

### What this study adds

The review highlights key recommendations (Figure 2) which, if implemented, have the potential to improve patient and carer experiences of frailty. Many of these needs could be addressed by incorporating palliative care services or approaches into care delivery throughout the trajectory of illness. As the recommendations arise from the synthesis of an international body of literature, they are relevant to an international audience.

**Figure 2:**
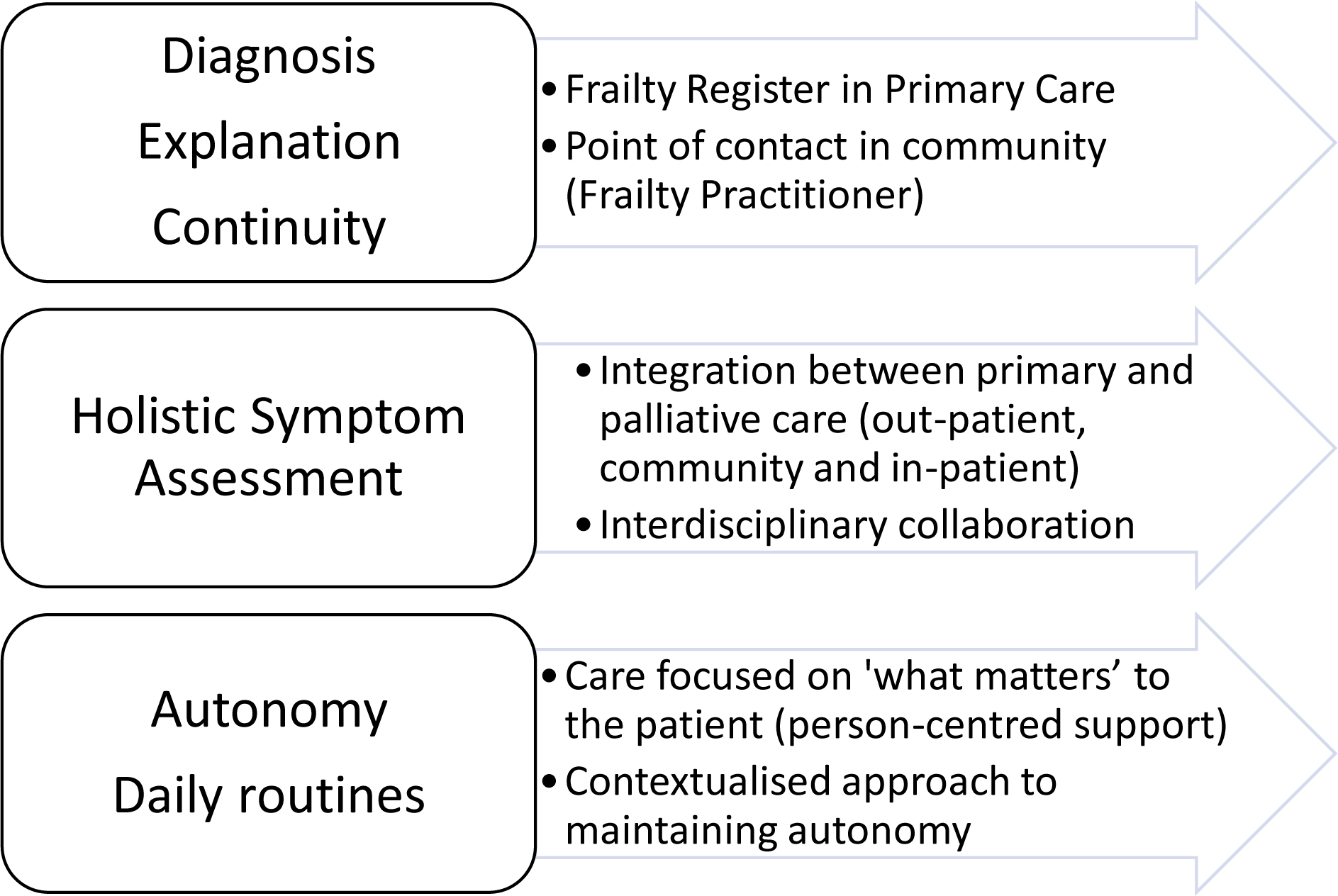
Identified themes and matched needs.

## Conclusion

This review has identified considerable literature exploring care experiences and needs of people with frailty and their relatives, and the experiences of care providers. People with frailty experience significant physical, emotional and social morbidity at the end-of-life, which current care providers are ill equipped to deal with. Acknowledging, these experiences and the currently unmet need for holistic palliative care is fundamental to configuring future policy and palliative care services. People with frailty and the healthcare professionals caring for them would benefit from greater awareness of, and involvement with, palliative care services.

## Data Availability

All data produced in the present study are available upon reasonable request to the authors

## Acknowledgements

Erika Gavillet (Medical Librarian at Newcastle University) assisted in the development of our search strategy

## Authorship

**EW, BB, DS, BH, FD contributed to the concept and design of the work and the analysis and interpretation of data; HO’K, EW, BB, DS, FD, LR, GS contributed to the acquisition of data; EW (or BH? DS?) led, and * contributed to, the analysis and interpretation of data; all authors contributed to the drafting of the article, revised it critically for important intellectual content and approved the version to be published; all authors have participated sufficiently in the work to take public responsibility for appropriate portions of the content.

## Data management and sharing

Search strategies and filters sufficient to replicate our searches are available as supplemental materials. All papers included in this review are referenced and available via their respective journals.

## Ethical approval

No ethical approval was required for this research, as the study was a review of existing published articles with no new primary data collected.

